# The effect of a video intervention on knowledge, awareness and perception of Natural Procreative Technology (NaProTechnology) among pharmacy undergraduate students in Nigeria

**DOI:** 10.1101/2023.05.29.23290690

**Authors:** Adaobi Uchenna Mosanya, Eziamaka Pauline Ezenkwele, Fausta Chioma Jacinta Emegoakor, Mmaduabuchi Okeh, Abdulmuminu Isah

## Abstract

**Background:** Natural Procreative Technology (NaProTechnology) is a system of management of infertility and other reproductive health issues which requires the application of a woman’s observation and record of key events throughout her menstrual cycle.

**Objectives:** The study assessed the knowledge, awareness, and perception of NaProTechnology as well as the effect of an educational intervention among pharmacy undergraduate students at University of Nigeria, Nsukka.

**Methods:** It was a cross-sectional, questionnaire-based study. The ethical approval was obtained from the research ethics committee of the Faculty of Pharmaceutical Sciences, University of Nigeria, Nsukka. At baseline, the knowledge, awareness and perception of the students were assessed. Followed by the administration of an educational video on NaProTechnology. Then a post intervention survey was done to assess the effect of the educational intervention.

**Key findings:** There were 410 and 350 students in the pre- and post-intervention surveys respectively with relatively equal number of males and females. Majority were between 18 and 29 years old. Less than 5% were married while the highest proportion of the respondents were from 300 level The knowledge, awareness, and positive perception of NaProTechnology among the pharmacy students prior to intervention were poor but improved markedly post intervention (P < 0.001).

**Conclusion:** A video intervention was effective in improving the short-term knowledge, awareness and positive perception of NaProTechnology among pharmacy students.

**Key messages:** *What is already known on this topic?:* Natural Procreative Technology (NaProTechnology) is a system of management of infertility based on the observation and record of fertility biomarkers by a woman.

*What this study adds?:* The level of knowledge, awareness, and positive perception of NaProTechnology among the pharmacy undergraduates students were poor prior to a video intervention but improved markedly afterwards

*How this study might affect research, practice or policy?:* This study would lead to the public health system approach of informing, educating and communicating NaProTechnology to pharmacy students to achieve long-term effects by using well-designed courses and curriculum implementation.

## Introduction

Natural Procreative Technology (NaProTechnology) is a system of management of infertility and other reproductive health issues which requires the application of a woman’s observation and record of key events throughout her menstrual cycle (Tham et al., 2012). These fertility biomarkers are noted following the principles of Creighton Model FertilityCare System (Stanford et al., 2008). The Creighton Model System (CrMS) is a natural method of fertility management based on the biomarkers observed in a woman’s cycle which indicate whether a woman is fertile or not (Hilgers, 2004). An important biomarker used in this system is the presence or absence of cervical mucus (Fehring, 2002).

NaProTechnology has diverse applications in the different aspects of a woman’s health especially in her reproductive life. Some of the gynaecological illnesses that can be treated using NaProTechnology are infertility, recurrent miscarriages, pre-menstrual syndrome, polycystic ovarian disease, menstrual irregularity, hormonal imbalances, ovarian cysts and post-partum depression (Tham et al., 2012). It is a model of health management with advantages because it forms the basis for the timing of specific diagnostic tests as well as the therapy and its monitoring (Hilgers, 2020). Through this method couples are empowered to become active and informed participants of their management (Camacho, 2018). This is quite different from assisted reproductive technology (ART) which is only aimed at achieving conception thereby leaving the couple still infertile afterwards because the root cause(s) of infertility were not treated (Keefe et al., 2012).

The NaProTechnologist is the medical doctor who has been trained to use the CrMS recordings to carry out a diagnostic evaluation and create a treatment regimen for each patient. The patient’s CrMS chart is evaluated by the NaProTechnologist on a regular basis to monitor treatment response. They also provide effective surgical gynaecologic care with the aim of restoring the reproductive system. The FertilityCare Practitioner (FCP) teaches the women how to chart their menstrual cycle according to the Creighton Model System. Moreover, pharmacists are also trained to become Creighton Model System Pharmacy Consultants as well as Fertility Care Practitioners (*Straight Talk on women’s health and fertility*, 2011). The Creighton Model System Pharmacy Consultants are expected to be knowledgeable in the science of NaProTechnology with respect to the pharmacological treatments, including bioidentical hormones and non-hormonal treatments (Hilgers, 2004). They compound the bioidentical hormones which are used in treating patients. Drugs like Vitamin B6, clomiphene citrate and low-dose naltrexone have also been found very useful in NaProTechnology treatment.

Over the years studies have been done regarding the effect of video intervention on fertility knowledge among university students (Conceição et al., 2017) among partnered women (Pedro et al., 2022), knowledge about infertility (Thomson et al., 2016). Nevertheless there is none assessing the effect of a video intervention on knowledge, awareness and perception of Natural procreative technology, a method of infertility management. In addition, because of the role that pharmacists play in NaProTechnology, it is pertinent to carry out this study among pharmacy undergraduate students.

## Objectives

Therefore, the purpose of this study was to assess the knowledge, awareness and perception of NaProTechnology as well as the effect of an educational intervention among pharmacy undergraduate students of University of Nigeria, Nsukka.

## Methods

### Study Design

A cross-sectional survey and an interventional study was conducted among pharmacy students at the University of Nigeria Nsukka, Enugu state in May 2021. The study was in two phases. The first phase was a baseline survey assessing the Knowledge, awareness and perception of the students regarding Naprotechnology. Then an eighteen-minute video on the overview of NaProTechnology was prepared by a gynaecologist and also an expert in NaProTechnology.

### Study Setting

The Faculty of Pharmaceutical Sciences began as a Department in the Faculty of Science in 1967. It was later raised to the status of a Faculty in 1980. At the time of study, the faculty was running a 5-year B-Pharm programme. The student population from 2^nd^ year to 5^th^ year in 2021 was 1531.

### Participants

All pharmacy students from 200 level to 500 level at the University of Nigeria Nsukka were eligible to participate in the study. All the students who were present at the time of the questionnaire distribution were invited to participate in the survey. A total of 410 students gave their consent to participate in the survey. A brief introduction was given about the investigator and their oral consent was received individually before administering the questionnaire.

### Variables

The correct responses for each question were reported as percentages. The total score of all the questions that made up the three domains of knowledge, awareness and perception were obtained. The Shapiro-Wilk tests of normality showed that the data were not normally distributed so the non-parametric methods for the inferential statistics were used. The median score served as the cut off point for the categorization of respondents into having either poor or good knowledge, good awareness or poor as well as positive or negative perception. Chi-square analysis was done to determine the association between the sociodemographic characteristics of the respondents and their level of knowledge, awareness and perception. The statistical significance for all the inferential analysis were taken at p < 0.05.

### Study size

A sample size of 308 was calculated using Raosoft sample size calculator (*Sample Size Calculator by Raosoft, Inc*., 2004). The Margin of error was set at 5% within a 95% confidence interval.

### Data sources

A seventeen item questionnaire was used as the data collection tool. It was structured into four sections: the first section consists of questions related to the sociodemographic characteristics of the respondents. The second section comprised of five knowledge questions. The third section were five awareness questions. While the last section were seven questions about their perception of NaproTechnology. The questionnaire was content and face validated by two experts in Clinical Pharmacy and a consultant gynaecologist. The final version was piloted among students who were excluded from participating in the final survey. The reliability test was done and a Cronbach alpha of 0.94 was obtained. The survey was done in two phases, the pre intervention phase and the post intervention phase. After the first phase, the video containing the information about NaProTechnology was posted on the WhatsApp platform of each of the classes. There were daily reminders from the investigator for the students to watch the videos. This lasted for one week. Then the post intervention survey was done by administering the questionnaires to only those who watched the video.

### Statistical Methods

The data was coded, entered into Excel spread, cleaned, exported to and analysed using Statistical Package for Social Sciences (SPSS) version 25. The statistical significance for all the inferential analysis were taken at p < 0.05.

### Ethics

The study was carried out according to the World Medical Association Declaration of Helsinki’s principles of research ethics (The World Medical Association Inc., 2013). Therefore, before beginning the study, the ethical clearance was sought and received from the Ethics Research Committee of the Faculty of Pharmaceutical Sciences, University of Nigeria Nsukka on 15^th^ July, 2020 (FPSRE/UNN/20/0007). In addition, an introduction of the study was given to each prospective participant and their informed consent was freely granted before they entered into research. Every measure was put in place to protect the privacy of the respondents and confidentiality of their personal data throughout the data collection and thereafter. It was also made known to the respondents that they were free to withdraw anytime from the study.

## Results

The calculated minimum sample size was 308. However, 410 students consented to participate in the study (Table 1). Moreover, in the post intervention phase, only 350 (Table 1) returned for the data collection. Based on the number of participants in the pre-intervention phase, the response rate was approximately 85%. The sociodemographic data were presented as frequencies and percentages.

**Table 1:** Sociodemographic characteristics of the respondents.

| Independent Variables | Pre intervention (n=410) | Post Intervention (n=350) |
| --- | --- | --- |
| **Sex** |  |  |
| Female | 207 (50.5) | 178 (50.9) |
| Male | 203 (49.5) | 172 (49.1) |
| **Age (in years)** |  |  |
| Less than 18 | 1 (0.2) | 1 (0.3) |
| 18-24 | 281 (68.5) | 235 (67.1) |
| 25-29 | 120 (29.2) | 108 (30.9) |
| 30-35 | 6 (1.5) | 5 (1.4) |
| Above 35 | 2 (0.5) | 1 (0.3) |
| **Marital Status** |  |  |
| Married | 17 (4.1) | 14 (4.0) |
| Single | 390 (95.1) | 334 (95.4) |
| Others | 3 (0.7) | 2 (0.6) |
| **Year of Study** |  |  |
| 2<sup>nd</sup> year | 73 (17.8) | 66 (18.9) |
| 3<sup>rd</sup> year | 181 (44.1) | 152 (43.4) |
| 4<sup>th</sup> year | 64 (15.6) | 47 (13.4) |
| 5<sup>th</sup> year | 92 (22.4) | 85 (24.3) |

A total of 410 students participated in the survey with relatively equal number of males and females. Majority of them were between 18 and 29 years old. Less than 5% were married while the highest proportion of the respondents were from 300 level.

The percentage of the respondents who gave correct answers to the knowledge questions before and after the educational intervention are shown in Table 2. The difference in the proportion of those who answered correctly the questions at baseline and those after the intervention was statistically significant. For the pre-intervention phase, the question that had the highest percentage (20%) of correct answer was “Is NaProTechnology harmful to human life?” For all the questions, there was a statistically significant difference between the pre and the post intervention responses. In addition, the percentage of the respondents who answered correctly the post intervention phase knowledge question “What is it used for?” was the highest (81.4%) for all the knowledge question. While the question, “can NaProTechnology be used in children?” received the lowest proportion of correct answers in the post intervention phase of the survey.

**Table 2:** Frequency and percentage of correct responses to the Knowledge questions.

| **Knowledge Questions** | **Pre-intervention**<br>**(n=410)** | **Post-intervention**<br>**(n=350)** | **p-value** |
| --- | --- | --- | --- |
| What is the full name of NaProTechnology? | **57 (13.9)** | **284 (81.1)** | **< 0.0001** |
| What is NaProTechnology used for? | **45 (11.0)** | **285 (81.4)** | **< 0.0001** |
| Who can use NaProTechnology? | **45 (11.0)** | **262 (74.9)** | **< 0.0001** |
| Is NaProTechnology harmful to human life? | **82 (20.0)** | **268 (76.6)** | **< 0.0001** |
| Can NaProTechnology be used in children? | **41 (10.0)** | **225 (64.3)** | **< 0.0001** |

The frequency and percentages of the correct responses to the awareness questions in the pre and post intervention surveys were presented in Table 3. The proportion of those who answered correctly the awareness questions were higher for most questions than those of the knowledge questions. The question that had the highest correct responses in both the pre and post intervention phases was “Are pharmacists involved in NaProTechnology?” and the percentages were 27.6 % and 84.6% respectively. There was also a statistically significant difference between the pre and the post – intervention responses. The awareness question with the lowest proportion of positive responses was “Have you ever heard of NaProTechnology?” However, in the post intervention phase, the question with the lowest proportion of correct responses was “Are there experts of NaProTechnology in Nigeria?”

**Table 3:** Frequency and percentage of correct responses to the Awareness questions.

| **Awareness Questions** | **Pre-intervention**<br>**(n=410)** | **Post-intervention**<br>**(n=350)** | **p-value** |
| --- | --- | --- | --- |
| Have you ever heard of NaProTechnology? | **50 (12.2)** | **278 (79.4)** | **< 0.0001** |
| Are there experts of NaProTechnology in Nigeria? | **91 (22.2)** | **236 (67.4)** | **< 0.0001** |
| Does NaProTechnology adopt individualized care? | **102 (24.9)** | **280 (80.0)** | **< 0.0001** |
| Are pharmacists involved in NaProTechnology? | **113 (27.6)** | **296 (84.6)** | **< 0.0001** |
| Are medications sometimes needed for NaProTechnology | 111 (27.1) | 289 (82.6) | < 0.0001 |

The correct responses to the perception questions are presented in Table 4. The question ‘Are there religious barriers that may impede the nationwide deployment of the service?” received the highest number of responses in the affirmative (70.7%) during the pre-intervention survey. While in the post-intervention phase, the question “Would the knowledge gained be useful to you in your future career as a pharmacist?” had the highest number of positive response (91.4%). There was also a statistically significant difference between the pre and post intervention responses.

**Table 4:** Frequency and percentage of positive responses to the Perception questions.

| Perception Question | Pre-intervention<br>(n=410) | Post intervention<br>(n=350) | p-value |
| --- | --- | --- | --- |
| Would knowledge gained be useful to you in your future career as a pharmacist? | 155 (37.8) | 320 (91.4) | < 0.0001 |
| If in need of NaProTechnology would you opt for it? | 138 (33.7) | 308 (88.0) | < 0.0001 |
| Would you be willing to pay for the services if in need of it? | 147 (35.9) | 311 (88.9) | < 0.0001 |
| Would you recommend it for family and friends in need of it? | 138 (33.7) | 306 (87.4) | < 0.0001 |
| Would you recommend it for clients in need of it in the future | 132 (32.2) | 304 (86.9) | < 0.0001 |
| Should it be deployed nationwide? | 142 (34.6) | 300 (85.7) | < 0.0001 |
| Are there religious barriers that may impede the nationwide deployment of the service? | 290 (70.7) | 93 (26.6) | < 0.0001 |

## Discussion

The knowledge, awareness, and positive perception of NaProTechnology among the pharmacy students prior to intervention were poor but improved markedly afterwards. This is understandable since NaProTechnology is a relatively new system of therapy and has not yet been included in the curriculum. It was developed in the 1980’s in the United States by Dr. TW Hilgers and his collaborators (Tham et al., 2012). The modal age group of the participants was between 18 and 29 years. This is expected because it seems to be the prevailing age bracket of undergraduates students in Nigeria (Aluh et al., 2019; Uleanya, 2021). Majority of the respondents were single. This is so because most undergraduate students are usually not ready for marriage. Educated women are less likely to be married at a younger age than uneducated women (Ononokpono et al., 2021). Probably before getting married they wish to have a stable employment with a steady income post graduation.

The brief video intervention showed a significant difference in their level of knowledge, awareness and perception. However, the percentages of the students with good knowledge, good awareness and positive perception were not 100% respectively. This may have been as a result of low level of engagement by some students. Learning outcome has been shown to be related to the level of engagemnt and learning goals (Seo et al., 2021). It is a also a fact that no matter how well a learning video is made, the level of assimilation depends on reflection and getting the processed information into the long term memory (Schulz & Iskru, 2021).

Is NaProTechnology harmful to human life? This question had the highest correct response rate of the knowledge section of the survey during the pre-intervention phase. This may have been as a result of intuition that NaProTechnology must be something safe for humans. In any case, NaProTechnology and the Creighton Model FertiltyCare System have been shown to be useful and harmless. Infact, couples have preferered it to assisted reproductive technology such as IVF because of its adavantages over the latter (Flores, 2016). While in the post intervention phase, the question “what is it used for?” received the highest percentage of correct answers. The uses of NaProTechnology had been mentioned earlier and they include management of hormonal imbalance, menstrual irregularity, polyccystic ovarian disease, recurrent miscarriages etc (Jemelka et al., 2013; Tham et al., 2012).

“Are pharmacists involved in NaProTechnology?” was the awareness question with the highest number of correct responses for both the pre and post interevention phases. Pharmacists are indispensable for the success of NaProTechnology. In fact there is a great demand for the natural form of progesterone, a hormone that has found its usefulness in the prevention of preterm delivery (Yadav et al., 2022). Pharamcists are needed to ensure that this naturally occuring hormone are comercially available in safe, accurate and sterile forms (Reed-Kane & Kirschbaum, 2006; Weber, 2005).

There was a high percentage of incorrect responses to the perception question: “Are there religious barriers that impede the nationwide depolyment of the service?” in the pre-intervention phase. However, in reality, there are no religoius bariers to the nationwide deployment of the service. The technology has been found to be ethically sound in line with the fundamental human right to life and respect for human dignity. NaProTechnology does not entertain abortion of fetuses because there is less likelihood of multiple pregnancies or need for hyperstimulation of the ovaries which carries with it high risk of maternal death (Vélez, 2012). Also it is not saddled with the prospect of freezing embryos or discarding some as does happen with in vitro fertilization (IVF) since NaProTechnology treatment encourages in vivo fertilization after treating the underlying cause (s) of infertility.

For the post intervention survey of the pharmacy students perception, one question emerged as most correctly answered; “Would the knowledge gained be useful to you in your future career as a pharmacist?”. Before the intervention, the correct response rate to the question above was low but increased afterwards. It shows clearly that the students percieved a possible career path for them in Naprotrechnology and would likely avail themselves of opportunities for training.

## Conclusion

In conclusion, there was a general poor knowledge and awareness in addition to a low positive perception of NaProTechnology among the surveyed students before the video intervention. However, the parameters of knowledge, awareness and perception improved remarkably after the video intervention which is an indication of a positive effect. The difference between the proportion of those who had good knowledge, good awareness and positive perception in the pre intervention phase and the post intervention phase was siginificant. There was a relatively higher awareness that pharmacists are needed in the delivery of NaProTechnology services.

## Data Availability

All data produced in the present study are available upon reasonable request to the authors

## Notes

**Declaration of Interest:** The authors declare that there are no conflicts of interest.

**Data availability:** The data supporting the findings are available from the corresponding author on reasonable request.

### Competing Interest Statement

The authors have declared no competing interest.

### Funding Statement

This study did not receive any funding

### Author Declarations

The ethical clearance was sought and received from the Ethics Research Committee of the Faculty of Pharmaceutical Sciences, University of Nigeria Nsukka on 15th July, 2020 (FPSRE/UNN/20/0007).

### Summary of Updates

Recommendation statements were removed frm the conclusion section of the abstract. Some sentences were removed from the conclusion of the main document. They were redundant.

## REFERENCES

Aluh, D. O., Chukwuobasi, T., & Mosanya, A. U. (2019). A Cross - Sectional Survey of Social Media Anxiety among Students of University of Nigeria. Journal of Mental Health and Human Behaviour, 24(1), 51–56. https://doi.org/10.4103/jmhhb.jmhhb

Camacho, A. (2018). The Effect of NaProTECHNOLOGY on Marital Interaction in Couples with Infertility. University of Connecticut.

Conceição, C., Pedro, J., & Martins, M. V. (2017). Effectiveness of a video intervention on fertility knowledge among university students: a randomised pre-test/post-test study. The European Journal of Contraception & Reproductive Health Care, 22(2).

Fehring, R. J. (2002). Accuracy of the peak day of cervical mucus as a biological marker of fertility. Contraception, 66(4), 231–235. https://doi.org/10.1016/S0010-7824(02)00355-4

Flores, R. L. (2016). Infertility in The Philippines and Natural Procreative (Napro) Technology : A Commentary. Scholars Academic Journal of Biosciences, 4(4), 328–331.

Hilgers, T. W. (2004). The Medical & Surgical Practice of NaProTECHNOLOGY. Pope Paul VI Institute Press.

Hilgers, T. W. (2020). The Identification of Postovulation Infertility with the Measurement of Early Luteal Phase (Peak Day +3) Progesterone Production. Linacre Quarterly, 87(1), 78–84. https://doi.org/10.1177/0024363919885551

Jemelka, B. E., Parker, D. W., & Mirkes, R. (2013). Virtual Mentor. American Medical Association Journal of Ethics, 15(3), 213–219.

Keefe, C. E., Mirkes, R., & Yeung, P. (2012). The evaluation and treatment of cervical factor infertility: A medical-moral analysis. Linacre Quarterly, 79(4), 409–425. https://doi.org/10.1179/002436312804827127

Ononokpono, D. N., Adebola, O. G., Gayawan, E., & Fagbamigbe, A. F. (2021). Modelling determinants of geographical Patterns in the Marital Statuses of Women in Nigeria. Spatial Demography, 0123456789. https://doi.org/10.1007/s40980-020-00072-5

Pedro, J., Fernandes, J., Barros, A., Xavier, P., Almeida, V., Costa, M. E., Schmidt, L., & Martins, M. V. (2022). Effectiveness of a video-based education on fertility awareness: a randomized controlled trial with partnered women. Human Fertility, 25(3), 522–533. https://doi.org/10.1080/14647273.2020.1854482

Reed-Kane, D., & Kirschbaum, K. (2006). Prevention of Preterm Delivery with Compounded 17a-Hydroxyprogesterone Caproate. International Journal of Pharmaceutical Compounding, 10(3), 165–171. https://doi.org/10.1056/nejmc072557

Sample Size Calculator by Raosoft, Inc. (2004). http://www.raosoft.com/samplesize.html

Schulz, J., & Iskru, V. V. (2021). Video in Education From ‘Sage on the Stage’’ to “TV Talk Show Host’’: Where to Next?”‘ Eurasia Journal of Mathematics, Science and Technology Education, 17(9), 1–6. https://doi.org/10.29333/ejmste/11154

Seo, K., Dodson, S., Harandi, N. M., Roberson, N., Fels, S., & Roll, I. (2021). Active learning with online video: The impact of learning context on engagement. Computers and Education, 165(December 2019), 104132. https://doi.org/10.1016/j.compedu.2021.104132

Stanford, J., Parnell, T., & Boyle, P. (2008). Outcomes From Treatment of Infertility WithNatural Procreative Technology in an Irish General Practice. The Journal of the American Board of Family Medicine, 21(5), 375–384.

Straight Talk on women’s health and fertility. (2011). Pope Paul VI Institute. http://www.drhilgers.com/tag/naprotechnology/

Tham, E., Schliep, K., & Stanford, J. (2012). Natural procreative technology for infertility and recurrent miscarriage: Outcomes in a Canadian family practice. Canadian Family Physician, 58(5), 267–274.

The World Medical Association Inc. (2013). World Medical Association Declaration of Helsinki: ethical principles for medical research involving human subjects. Journal of the American Medical Association, 310(20), 2191–2194. https://doi.org/10.1093/acprof:oso/9780199241323.003.0025

Thomson, A. A., Brown, M., Zhang, S., Stern, E., Hahn, P. M., & Reid, R. L. (2016). Evaluating Acquisition of Knowledge about Infertility Using a Whiteboard Video. Journal of Obstetrics and Gynaecology Canada, 38(7), 646–650. https://doi.org/https://doi.org/10.1016/j.jogc.2016.03.010

Uleanya, C. (2021). Exploring Undergraduates’ Perception on Assessments and Feedbacks at Selected Nigerian and South African Rural Universities. Universal Journal of Educational Research, 9(4), 836–843. https://doi.org/10.13189/ujer.2021.090417

Vélez, J. (2012). An Ethical Comparison between In-Vitro Fertilization and NaProTechnology. The Linacre Quarterly, 79(1), 57–72. https://doi.org/10.1179/002436312803571465

Weber, C. (2005). AGENTS OF CHANGE. The Catholic World Report, 15(4), 38–47.

Yadav, G., Gupta, S., Singh, P., Kansara, M., Kathuria, P., Gothwal, M., & Sharma, C. (2022). The role of vaginal progesterone in established pre - term labor : A randomized controlled trial. https://doi.org/10.4103/jfmpc.jfmpc

